# “Let’s better not choose someone with HIV”: Dutch professionals’ attitudes and job promotion decisions regarding candidates living with HIV and the mediating role of employability concerns

**DOI:** 10.1101/2024.08.21.24312366

**Authors:** Marco Gaetani, Karen Schelleman-Offermans, Kai J. Jonas

**Author notes:** Correspondence concerning this article should be addressed to Kai J. Jonas, Department of Work and Social Psychology, Maastricht University, Maastricht, the Netherlands. **Author note** The authors would like to thank Martijn Bruil and the members of the community advisory group for their critical input and support. KJJ received funding from Gilead and ViiV Healthcare unrelated to the topic of this study. The authors have no conflicts of interest to disclose. Mindful that our identities can influence our approach to science and our view on the topic, the authors wish to provide the reader with information about our backgrounds. With respect to gender identity and sexual orientation, two authors self-identified as cisgender gay men and one as a cisgender heterosexual woman. With respect to ethnicity, all authors self-identified as white. To include the views of professionals living with HIV, community members were involved in the design and initiation of this research project. Data used will be made available upon reasonable request to the corresponding author.

## Abstract

Despite advancements in HIV health management, workplace stigma against people living with HIV (PLHIV) prevails. However, current literature has focused on labor market access rather than job promotion for PLHIV. This study investigated how professionals living with HIV are perceived compared to those with other chronic health conditions, and explored how employability concerns mediate the association between stigmatizing attitudes and discriminatory practices against PLHIV in job promotions.

A nationally representative sample of 640 heterosexual and HIV-negative Dutch professionals completed an online questionnaire designed with input from professionals living with HIV for a cross-sectional study. Data was analyzed using multidimensional unfolding (MDU) to uncover latent dimensions driving participants’ perceptions of PLHIV and structural equation modeling (SEM) to test the mediating role of employability concerns.

The MDU analysis revealed that participants held similar perceptions of and attitudes towards job promotion applicants living with HIV and applicants surviving cancer, compared to those living with other chronic conditions. The SEM showed that prejudice and support for discriminatory policies were significantly associated with discriminatory behavior towards PLHIV, while stereotypes and social distancing attitudes were not. Employability concerns partially and fully mediated, respectively, the relationship between prejudice and support for discriminatory policies, and discriminatory decision-making.

Consistent with previous research on discriminatory hiring practices, our findings highlight the crucial role of employability concerns in driving job promotion discrimination against PLHIV and the need to address knowledge gaps about HIV as a manageable condition among Dutch professionals as a means to combat stigma and safeguard careers of PLHIV.

## 1. Introduction

HIV treatment and the introduction of combination antiretroviral therapy have transformed HIV infection into a manageable and noncontagious chronic condition, resulting in a near-normal life expectancy for people living with HIV (PLHIV) (Deeks et al., 2013; Nakagawa et al., 2013). Despite these advances, HIV remains highly stigmatized worldwide, even in countries with advanced healthcare systems and robust human rights protections, such as the Netherlands (Stutterheim et al., 2014). People living with HIV often face rejection and unequal treatment in multiple life domains, including the workplace (Bruil & Jonas, 2016; Chan et al., 2020). Several studies conducted in the Global South indicated that, despite the existence of formal national non-discrimination policies, people living with HIV still experience concerningly high levels of workplace discrimination, including exclusion in the hiring process, forced disclosure of their HIV status, and job loss (e.g., Deane et al., 2022; GNP+, 2018; Ho & Goh, 2017; Sprague et al., 2011; Twinomugisha et al., 2020). Although further investigation and efforts to tackle discrimination in job access and maintenance are crucial, job hiring or termination decisions are not the only factors impacting employment and career progression for professionals living with HIV (Dalgin, 2018). Findings from the HIV Stigma Index revealed significant percentages of individuals living with HIV experiencing changes in job descriptions, alterations in work nature, or denial of promotions due to their HIV status, with discrimination, rather than ill health, being the leading cause (GNP+, 2018). These findings indicate that HIV is still a major obstacle not only to employment security but also to job advancement and the quality of working life for professionals living with HIV. Furthermore, statutory protections against overt discriminatory hiring practices for individuals living with chronic conditions have likely led to subtler forms of discrimination against them, particularly in regard to their career trajectories and advancement opportunities (Fisher & Henrickson, 2019). However, despite extensive literature on labour market access or labor force participation (e.g., Maulsby et al., 2020; Nachega et al., 2015; Rajabiun et al., 2023), research on the role of HIV-related stigma in shaping career advancement opportunities for PLHIV in high-income countries such as the Netherlands is limited. With more than 24,000 people living with HIV in the Netherlands (van Sighem et al., 2023) and one in three of them estimated to encounter workplace discrimination (Stutterheim et al., 2022), it is essential to continue investigating how PLHIV are perceived and the impact of HIV stigma on career advancement.

Tackling HIV stigma is crucial not only due to its negative impact on prevention, treatment, and support initiatives (Stangl et al., 2012), but also because it perpetuates unfounded concerns about HIV as a limiting health condition impairing individuals’ ability to live and sustain their job (Janssens et al., 2021; Vornholt et al., 2013). For instance, previous research (Corrigan et al., 2000; Toppenberg et al., 2015) suggests that HIV could still be perceived by poorly informed populations as more similar to severe and life-threatening health conditions (e.g., cancer) than others (e.g., physical disability, depression). Similarly, a global survey conducted across 50 low, middle, and high income countries found that 35% of respondents believed that people living with HIV should not be allowed to work directly with those who do not have HIV, 63% of whom believed that people with HIV cannot be productive at work because of their ill health (International Labour Organization, 2021). Prior research documented the role played by these inaccurate preconceptions about the work ability, job performance, and health status of candidates living with HIV in deterring employers’ decision to interview or hire them (e.g., Liu et al., 2012; Rao et al., 2008). This suggests that also career-advancement discrimination towards people living with HIV could be driven by concerns about their (sustainable) employability, namely about the possibility for them to carry out their job and function in the labor market with satisfactory levels of, among others, performance, health, and well-being (Fleuren et al., 2020). However, these aspects have not been extensively investigated in relation to job promotion of professionals living with HIV.

Building on the empirical gap in the exploration of career trajectories among people living with HIV, this study utilized the widely recognized Earnshaw and Chaudoir’s HIV Stigma Framework (2009) to gain insight into how stigmatizing attitudes and concerns contribute to shaping the career trajectories of professionals living with HIV. According to this framework, individuals not living with HIV exhibit their stigma towards PLHIV through prejudice (i.e., negative emotions and feelings towards PLHIV, including disgust, anger, shame, and fear), stereotypes (i.e., potentially inaccurate group-based beliefs about PLHIV applied to specific PLHIV), and discrimination (i.e., behavioral expressions of prejudice towards PLHIV, encompassing social distancing and support for discriminatory social policy against them), in line with a tripartite model of attitude.

### 1.1 The Present Study

The present research primarily sought to examine Dutch professionals’ perceptions of HIV compared to other chronic conditions in relation to job promotion. Additionally, this study aimed at investigating whether stigmatizing attitudes (i.e., prejudice, stereotypes, and discrimination; Earnshaw & Chaudoir, 2009) towards professionals living with HIV are associated with a higher likelihood of discriminating against them in job promotion contexts, with this relationship being mediated by concerns about their employability. Therefore, the three research questions guiding the current study are:

1. *How do professionals perceive job promotion candidates living with HIV compared to those living with other chronic conditions (e.g., depression, hypertension)?*
2. *Are stigmatizing attitudes (i.e., prejudice, stereotypes, discrimination) towards people living with HIV associated with professionals’ tendency to engage in job promotion discrimination against them?*
3. *Is the association between stigmatizing attitudes and engagement in job promotion discrimination mediated by professionals’ concerns about the employability of people living with HIV?*

## 2. Methods

### 2.1 Study design and population

Cross-sectional data was collected in late 2021 via an online questionnaire administered in Dutch to a nationally representative sample of heterosexual and HIV-negative Dutch professionals, following approval from the Ethics Committee of Maastricht University, Faculty of Psychology and Neuroscience (ERCPN: 188_11_02_2018_S20). The administered questionnaire was designed using a community-based participatory approach (Holkup et al., 2004), entailing a collaboration between community members (i.e., professionals living with HIV) and researchers throughout the whole formulation of the questionnaire items. A community advisory group consisting of 10 professionals living with HIV provided input through focus group discussions and individual interviews, with a subset of this group contributing to designing the final survey items. A final sample of 640 heterosexual participants representative of the Dutch workforce at the time of data collection completed the questionnaire. Being representative of the Dutch workforce at the time of data collection, the majority of the sample comprised of men, atheists, and highly educated participants, with a mean age of 41.72 years (*SD* = 11.43, *range*: 19-67) (Table 1). Constructs and socio-demographic variables were measured in the order in which they are reported in section 2.2.

**Table 1.** Respondents’ socio-demographic characteristics. *Note*: percentages are estimated using sample weights.

| Sample | $N = 640$ |
| --- | --- |
| Gender |  |
| Woman | 44% |
| Man | 56% |
| Age |  |
| 18-24 | 5.8% |
| 25-34 | 27% |
| 35-44 | 26% |
| 45-54 | 26% |
| 55-64 | 13% |
| > 65 | 1% |
| Faith |  |
| Atheist | 53% |
| Religious | 47% |
| Education |  |
| Primary education | 2% |
| Lower secondary education | 16% |
| Upper secondary education | 38% |
| Higher education | 43% |
| Geographical residence |  |
| Border municipalities | 13% |
| Randstad (Amsterdam, Rotterdam, Utrecht, The Hague) | 31% |
| West Netherlands | 8% |
| North Netherlands | 20% |
| East Netherlands | 24% |
| South Netherlands | 4% |
| Company size |  |
| Self-employed/works independently | 8% |
| 1 to 15 employees | 13% |
| 16 to 100 employees | 20% |
| 100 to 500 employees | 21% |
| 500 to 5000 employees | 21% |
| More than 5000 employees | 17% |

### 2.2 Measures

*Promotion decision* was assessed through a single item asking participants to imagine having a say in determining who would get a hypothetical managerial position becoming available in their company (involving leading a team of 12 people, some overtime, client interactions, and 3 to 4 international trips per year). Participants had to rank six potential candidates, all equally qualified but with different chronic health conditions (i.e., HIV, hypertension, cancer one year ago, diabetes, Crohn’s disease, and depression), based on how suitable (1 = most suitable to 6 = least suitable) they believed each of them to be for this potential managerial position. This resulted in six variables (one for each health condition) ranging from 1 (if the participant believed the candidate with the health condition in question to be the most suitable for the managerial position) to 6 (least suitable compared to all the other conditions). While all of these six variables were employed in the multidimensional unfolding analysis, only the variable related to the candidate living with HIV was included as outcome in the mediation analysis, due to its relevance to the research question.

*Concerns about the employability of PLHIV* (α = .90) were measured by asking individuals to express their agreement (1 = completely disagree to 5 = completely agree) with six items, to gain insights into the beliefs driving their perceptions of suitability for job promotion of candidates living with HIV. Examples of items include “This person cannot fully cope with the challenges of the role”, “This person is likely to have too much absenteeism due to illness in the future”, “This person faces health risks during international travel”, and “This person would overly burden their team members due to their illness”.

*Prejudice against PLHIV* (α = .92) was evaluated by having participants rate their agreement with three items assessing negative emotions and feelings towards PLHIV (Earnshaw & Chaudoir, 2009) on a Likert scale from 1 (completely disagree) to 5 (completely agree). Examples of the items include “I feel nervous around people with HIV” and “I feel anxious around people with HIV”.

*Stereotypes about PLHIV* (α = .80) were measured via four items exploring participants’ potentially inaccurate group-based beliefs about PLHIV (Earnshaw & Chaudoir, 2009). Participants rated their agreement with statements such as “Most people with HIV are sex workers” and “Most people with HIV have had many different sexual partners” using a 5-point Likert scale (1 = completely disagree to 5 = completely agree).

*Discrimination against PLHIV* was assessed using items measuring two dimensions of the behavioral expression of prejudice directed at people living with HIV, namely social distancing and removal of rights (Earnshaw & Chaudoir, 2009). Specifically, *social distancing towards PLHIV* (α = .80) was evaluated by asking participants to indicate their level of agreement on a 5-point Likert scale (1 = completely disagree to 5 = completely agree) with five items, such as “People with HIV should be able to work together with others” (reverse scored), and “If I found out a family member has HIV, I would be willing to take care of them in my family” (reverse scored). *Removal of rights against PLHIV* (α = .66) was assessed using three items requiring individuals to express their agreement (1 = completely disagree to 5 = completely agree) with statements including “I believe it is justified that people with HIV have fewer career opportunities in organizations” and “I would find it acceptable if people with HIV were to pay a higher premium for their health insurance”.

*Covariates* used in the mediation analyses encompassed participants’ characteristics potentially associated with HIV stigma endorsement, in order to mitigate the influence of confounders and increase the precision of the estimates for parameters of interest. Dummy variables for nominal and ordinal variables with at least two categories were created. The background variables included were gender, educational level, age in years, company size, geographical residence, faith, social contact (i.e., knowing any person living with HIV), and HIV-related social desirability (measured via ten 5-point Likert items assessing the Impression Management (α = .88) and Self-Deception (α = .76) dimensions; examples of items include “I try not to show prejudices in my behavior towards people with HIV to avoid disapproval from other people” for the Impression Management subscale and “Due to my own conviction, I am motivated not to have prejudices towards people with HIV” for the Self-Deception subscale).

### 2.3 Data analysis

Statistical analyses were carried out in three steps. First, to gain insight into the perceptual map plausibly guiding participants’ preference judgments for candidates with various chronic health conditions to hold managerial roles, a multidimensional unfolding (MDU) solution (section 3.1) was computed using the smacof package (de Leeuw & Mair, 2009; Mair et al., 2022) in R, version 4.3.2 (R Core Team, 2023). MDU is a scaling technique used for the exploratory analysis of preference or ranking data, which allows to plot individuals (i.e., participants) and objects (e.g., chronic diseases) in a shared multidimensional space to uncover latent dimensions of participants’ judgments of the objects. Building upon the premise that participants share a common perception of the objects but differ in their preferences (Borg et al., 2018), MDU maps original preference data (also called “dissimilarities”) into distances between individuals and the objects they judged. In the context of this research, the closer a chronic condition is to a person’s point, the stronger their preference for promoting a candidate with that condition. There are three main variants of MDU models, each preserving different properties of the original data when computing distances. The first variant, ordinal MDU, transforms dissimilarities into distances that preserve the *order* of original preferences, namely ensuring that distances maintain the same order as dissimilarities. The second variant, interval MDU, transforms dissimilarities into distances by preserving the relative *differences* between object ratings in the original data, so that the differences between distances match those between dissimilarities. Finally, the third variant, ratio MDU, maps dissimilarities into distances by preserving their *ratios*, ensuring that the ratios between distances correspond to the ratios between dissimilarities. When it can be assumed that data are comparable across rows (i.e., participants), the transformation used to convert dissimilarities into distances can be applied uniformly across all rows (“unconditional” unfolding). However, if data comparability across rows cannot be assumed (e.g., due to participants’ response style artifacts), the researcher may opt to apply a person-specific transformation to each row (“row-conditional” unfolding). Although the latter approach can be desirable for certain theoretical reasons, it further reduces the constraints that the data impose on the distances in the final solution, thereby reducing the robustness of the findings (Borg et al., 2018). The choice of the MDU (ordinal, interval, or ratio) model depends on at least two aspects, namely the scale level of the original data and the desired balance between robustness and overfit of the solution (Borg et al., 2018). Regarding the first aspect, it generally makes sense to select a MDU model that corresponds to the scale level of the original preference data (e.g., ordinal MDU for ordinal data). For example, if the original data are ordinally scaled (as in this study), the rank-order of participants’ preferences may more accurately reflect their true cognitions, while differences between ratings might not correspond to any psychological quantities (Borg & Groenen, 2005), making an ordinal solution preferable to an interval or ratio one. The second aspect deals with the observation that interval and ratio MDU models are generally more robust than ordinal solutions, as they impose greater restrictions that reduce data overfitting, minimize the risk of degenerate solutions, and improve the robustness and replicability of results (Borg et al., 2018). Therefore, Borg and colleagues (2018) recommend starting the analysis with more robust models (e.g., ratio unconditional unfolding) and only resorting to weaker ones (e.g., ordinal row-conditional unfolding) in case the former cannot be salvaged. For this study, a two-dimensional unfolding solution using multiple random starts and an interval unconditional transformation of original ranking data was used, in line with recommendations from reference literature (Borg et al., 2018; Mair et al., 2016). The quality of the MDU solution was evaluated using a badness-of-fit measure called stress, which indicates the level of distortion in the visualization of the original preference data matrix (Dexter et al., 2018). In addition to stress, other goodness-of-fit tools were examined, including the initial configuration of the solution, the scree plot of stress values, the Shepard diagram of original data transformations, and the permutation test (Mair et al., 2016). This comprehensive evaluation was necessary because, although a high stress value is generally considered undesirable, it does not automatically warrant rejection of the solution, as an excellent fit may also indicate that more error is being reproduced (Borg et al., 2018). A two-dimensional solution was chosen to minimize the number of dimensions for ease of visualization and interpretation of the final configuration plot (Mair et al., 2016), supported by the analysis of scree plots (Figure S1), which revealed negligible differences in stress values between two and three-dimensional solutions. Furthermore, an interval transformation of participants’ preferences was chosen as the disparity in stress between the ordinal (stress = 0.27) and interval (stress = 0.33) solutions was not substantial (Figure S2), while a ratio transformation was deemed inadequate as the function linking preferences to distances did not intersect the origin, indicating that the data were mapped into distances that did not preserve their ratio (Borg et al., 2018). Results using an ordinal transformation of dissimilarities are also provided (Figure S3), showing no substantial differences from the results obtained with the interval solution.

In the second step, IBM SPSS Statistics, version 28 (IBM Corp., 2021) was used to assess the internal consistency and dimensionality of each scale included in the analysis, employing Cronbach’s α coefficient and exploratory factor analysis, respectively. Scale scores were then computed by summing the responses to each item. Descriptive univariate and bivariate statistics were subsequently conducted to evaluate the distribution of the variables of interest and to examine their associations (section 3.2).

In the third and last step, mediation analyses (section 3.3) were conducted using structural equation modelling in MPlus, version 7.3 (Muthén & Muthén, 1998). Prejudice, stereotypes, social distancing, and removal of rights were included as independent variables, concerns about employability as mediator, and promotion decision as dependent variable. Model parameters were estimated using maximum likelihood estimation, while bootstrapping was employed to estimate the coefficients of indirect effects. Multicollinearity among constructs was assessed by examining the Variance Inflation Factor, ensuring it did not exceed the conventional threshold of 5. Finally, there was no missing data.

## 3. Results

First, the multidimensional unfolding analysis is presented to explore how professionals included in the study perceived the suitability for promotion to leadership positions of candidates living with HIV compared to those with other chronic conditions. Next, descriptive statistics and correlations are reported to provide an initial snapshot of stigmatizing attitudes and discriminatory decisions against candidates living with HIV within the sample. Finally, structural equation modeling findings are presented to further investigate the relationship between stigmatizing attitudes and discriminatory job promotion decisions, and to examine the mediating role of employability concerns.

### 3.1 Perceptions of job promotion candidates with different chronic conditions

To dive into the perception of job promotion candidates with different conditions, an exploratory multidimensional unfolding analysis was conducted. The top side of Figure 1 illustrates the two-dimensional joint space compatible with participants’ judgments (permutation test with N_replications_ = 500, *p* < .001). The results suggested that individuals differentiated among candidates with various chronic conditions clustering them into three main groups (indicated by the proximity of objects in the shared plot): one comprising HIV and cancer, another comprising Crohn’s disease, diabetes, and hypertension, and a final group comprising depression only. A plausible interpretation for the two dimensions pertains to the aspects of perceived mortality risk or unresponsiveness to therapy (vertical dimension) and perceived controllability or mildness of symptoms experienced (horizontal dimension) for each chronic condition. Sticking to this interpretation, HIV and cancer would be perceived as the most life-threatening, in contrast to depression. Furthermore, individuals living with conditions such as Crohn, HIV, and depression would be seen as having the most uncontrollable or severe symptoms, as opposed to those dealing with hypertension or having overcome cancer one year ago.

**Figure 1.**
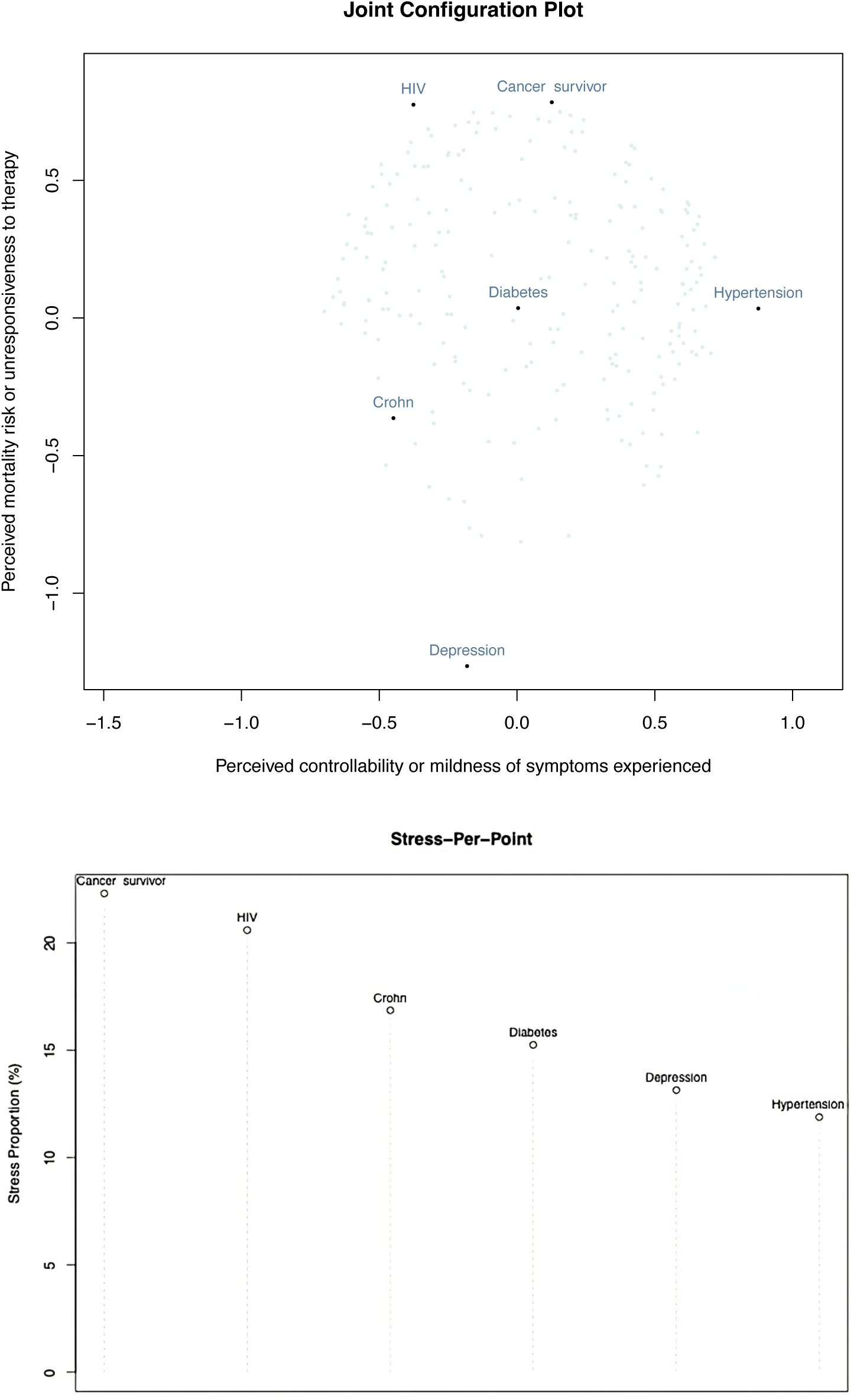
Top: Two-dimensional unfolding solution plot of individuals (light dots) and objects (dark dots). Bottom: Stress Per Point diagram illustrating the contribution of each object to the overall stress of the solution.

To better understand the contribution of each object (i.e., chronic condition) to the overall stress (i.e., misfit) of the chosen solution, a Stress-Per-Point diagram was generated (bottom graph in Figure 1). The graph suggests that cancer and HIV posed greater challenges for the algorithm in positioning them on the final plot, as indicated by their higher stress contribution proportion compared to other conditions such as hypertension and depression.

### 3.2 Association between stigmatizing attitudes and discriminatory promotion decision

To preliminarily assess the relationship between the main variables included in the analysis, descriptive statistics and correlations were conducted (Table 2). The means and standard deviations showed the sample to be rather socially desirable concerning HIV-related topics, moderately prejudiced and holding social distancing attitudes towards people living with HIV, and to have moderate levels of stereotypes, attitudes concerning the removal of rights for PLHIV, and concerns about their employability. Furthermore, the data revealed the perception of suitability (to handle a managerial position) of potential candidates living with HIV to be at medium levels, indicating an average preference for a candidate living with HIV over three other chronic conditions. This perception, however, varied significantly across participants, as indicated by the relatively high standard deviation.

**Table 2.**
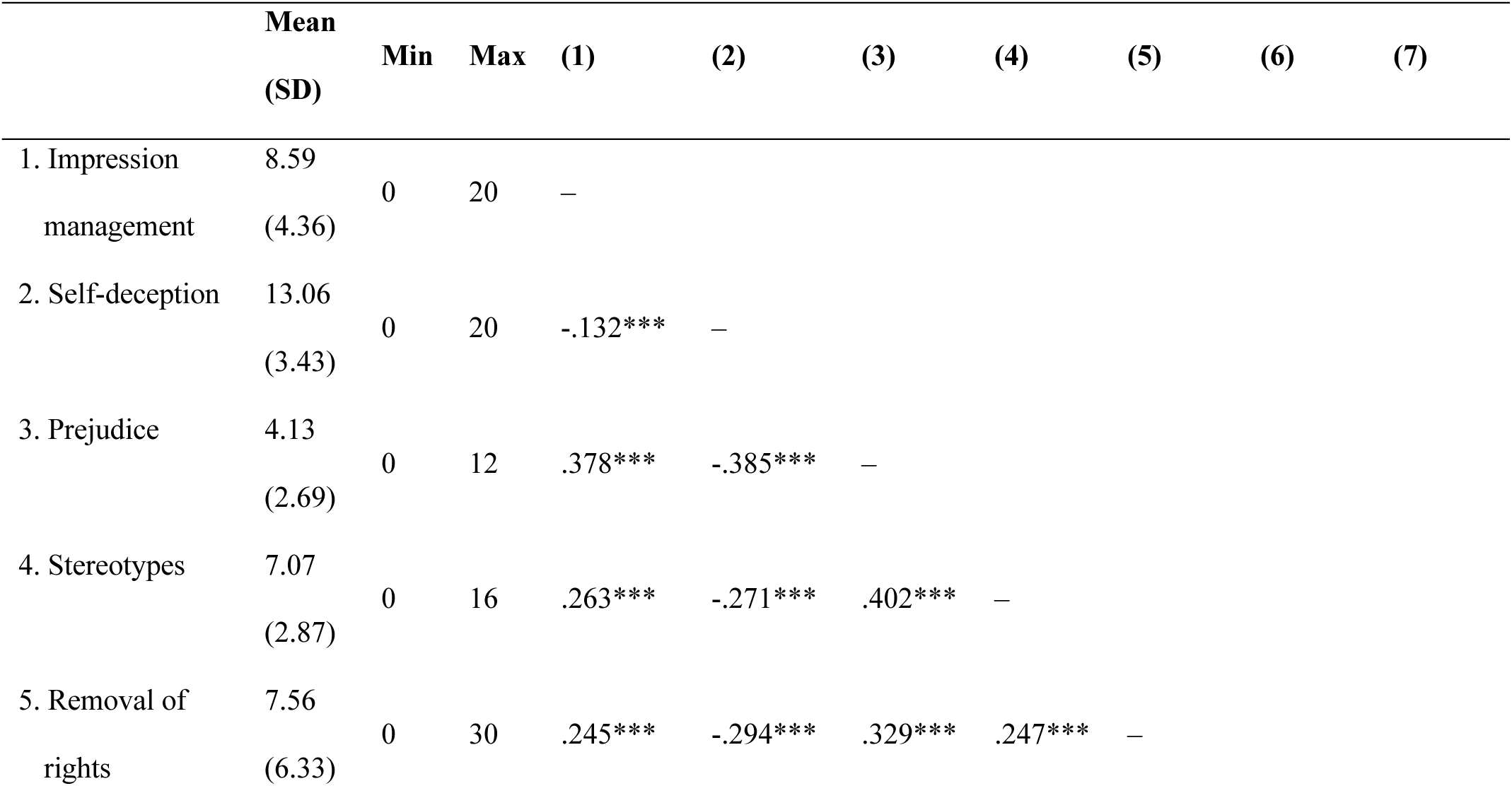
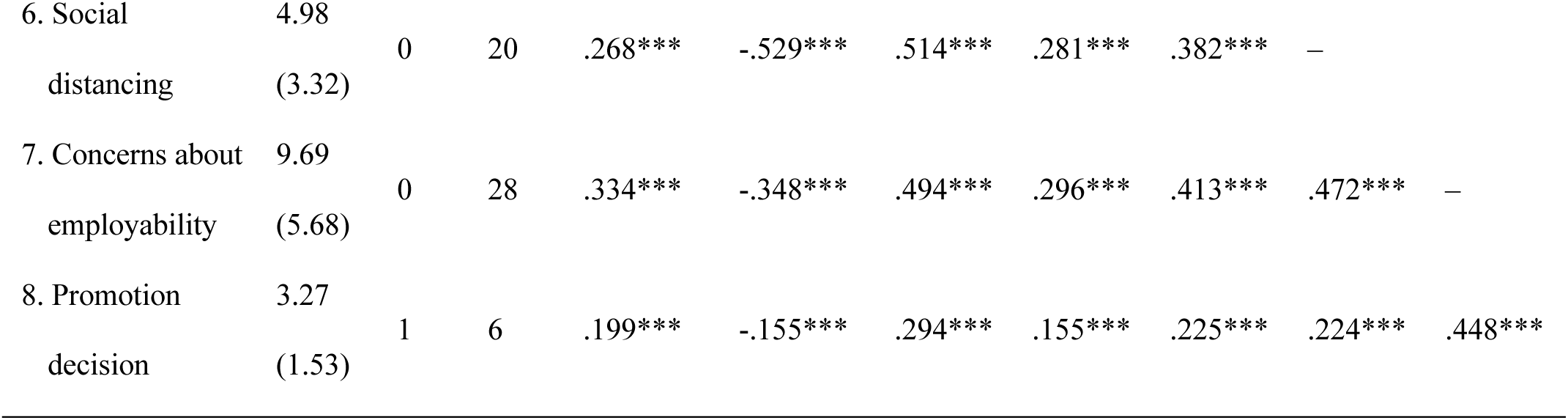
Scores description and bivariate correlations of the scales included in the analysis. *Note*: descriptive statistics are estimated using sample weights. *** p < 0.001, ** p < 0.01, * p < 0.05

Pearson correlations displayed on the right side of the table indicated all the main variables included in the mediation model to be (weakly to strongly) positively correlated. Individuals endorsing more stigmatizing attitudes towards PLHIV expressed greater concerns about their employability and tended to perceive them as less suitable for promotion to leadership positions compared to people with other chronic conditions.

### 3.3 Mediating role of employability concerns

To investigate the mediating role of employability concerns, mediation analysis was employed. Figure 2 and Table 3 summarize the standardized estimates from the structural equation model specified to explore the role of employability concerns in mediating the relationship between HIV-stigmatizing attitudes and job promotion decisions towards individuals living with HIV. The proportion of variance in the outcome explained by the entire model was R^2^ = .25, while that of the mediator was R^2^ = .35.

**Figure 2.**
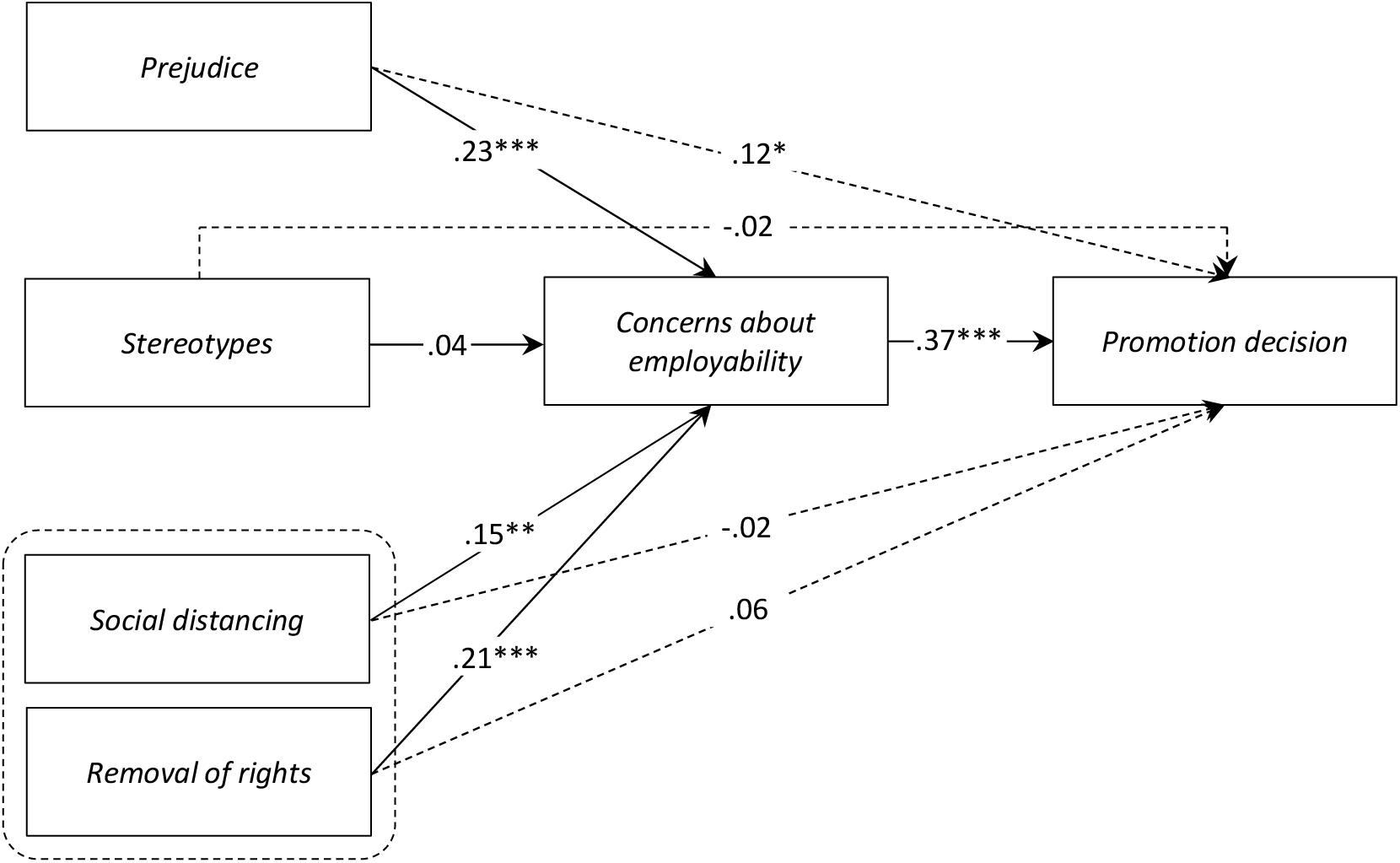
Structural equation model with prejudice, stereotypes, and discriminatory attitudes (i.e., social distancing and removal of rights) as exogenous variables, concerns about employability as mediator, and promotion decision as outcome. *Note*: standardized coefficients are reported; dashed lines indicate the direct effects of the exogenous variables on the outcome; effects were estimated net of participants’ gender, educational level, age, company size, geographical residence, faith, social contact, and HIV-related social desirability. *** p < 0.001 ** p < 0.01 * p < 0.05

**Table 3.** Total, direct, and indirect standardized effects of the exogenous variables on the outcome. *** p < 0.001 ** p < 0.01 * p < 0.05

| Parameter estimates | *B* (*SE*) | $\beta$ (95% CI) |
| --- | --- | --- |
| *Total effects* |  |  |
| Prejudice → Promotion decision | .12*** (.02) | .21*** (.11, .31) |
| Stereotypes → Promotion decision | -.00 (.02) | -.01 (-.09, .07) |
| Social distancing → Promotion decision | .01 (.02) | .03 (-.06, .13) |
| Removal of rights → Promotion decision | .03** (.01) | .13** (.05, .22) |
| *Direct effects* |  |  |
| Prejudice → Promotion decision | .07* (.02) | .12* (.02, .22) |
| Stereotypes → Promotion decision | -.01 (.02) | -.02 (-.11, .05) |
| Social distancing → Promotion decision | -.01 (.02) | -.02 (-.12, .07) |
| Removal of rights → Promotion decision | .01 (.01) | .06 (-.02, .14) |
| *Indirect effects* |  |  |
| Prejudice → Concerns → Promotion decision | .05*** (.01) | .08*** (.04, .13) |
| Stereotypes → Concerns → Promotion decision | .00 (.00) | .01 (-.01, .05) |
| Social distancing → Concerns → Promotion decision | .02** (.00) | .05** (.02, .09) |
| Removal of rights → Concerns → Promotion decision | .01*** (.00) | .07*** (.04, .11) |

The inspection of the total effects of the exogeneous variables on the outcome revealed that both prejudice and support for PLHIV rights removal were positively associated with participants’ tendency to act in a discriminatory way against candidates living with HIV, whereas stereotypes and social distancing attitudes were not. The indirect effects of both prejudice and removal of rights on promotion decision were positive and statistically significant; in particular, the association between the emotional (i.e., prejudice) and behavioral (i.e., removal of rights) components of attitude and discriminatory decision-making was partially and fully mediated, respectively, by concerns about the employability of candidates living with HIV. Thus, endorsing stigmatizing attitudes against PLHIV was associated with increased concerns about their employability, which, in turn, correlated with a greater tendency to discriminate against candidates living with HIV in job advancement decisions.

## 4. Discussion

The aims of this study were to (1) explore Dutch professionals’ perceptions of HIV compared to other chronic health conditions in relation to job promotion decisions, (2) examine whether stigmatizing attitudes towards professionals living with HIV were associated with participants’ tendency to engage in job promotion discrimination against them, and (3) assess the role of concerns about the employability of people living with HIV in mediating the relationship between the endorsement of discriminatory attitudes and engagement in discriminatory job promotion decisions.

In general, despite noteworthy levels of social desirability, participants exhibited a slight endorsement of prejudice and social distancing attitudes towards professionals living with HIV, alongside moderate levels of stereotypes, support for discriminatory policies against them, and concerns about their employability.

Regarding the first aim, results from the multidimensional unfolding analysis suggested that participants perceived applicants to a potential job promotion living with HIV not to be substantially different from those who survived cancer in terms of mortality or unresponsiveness to treatment of their health condition. The perceived similarity between these two conditions is reflected in the proximity of HIV and cancer in the perceptual map presented in Figure 1, as well as in their significant contribution to the overall misfit of the final solution, as shown in the Stress-Per-Point diagram. One explanation for the high stress contribution of these two conditions is that evaluating candidates with a history of cancer or living with HIV required a more complex cognitive process from participants, who applied different evaluation criteria or held weaker or ambivalent attitudes toward individuals with these conditions compared to those with other chronic conditions. Overall, the HIV-cancer cluster detected in the present study is consistent with previous evidence from immersive virtual environment technology by Toppenberg and colleagues (2015) showing that Dutch university students exhibited slower approach and quicker withdrawal behaviors towards individuals with HIV and cancer compared to those with a broken leg. In addition, the considerable distance of depression from all other conditions on the perceptual map also reflects prior evidence on mental health workplace stigma. For instance, Dalgin and Bellini (2008) showed that employers exhibit a preference for hiring individuals with sensory or physical disabilities over those with intellectual or psychiatric disabilities, rating the employability of applicants with psychiatric disabilities significantly lower.

Concerning the second research question, the structural equation model revealed that higher levels of prejudice and support for discriminatory policies positively predicted discriminatory behavior towards candidates living with HIV, while stereotypes and social distancing attitudes did not. In support of this finding, literature suggests that negative emotions toward minority groups are more predictive of rejecting and discriminatory behavior towards them than stereotypes (Thornicroft et al., 2007). For instance, emotional prejudice was shown to predict attitudes towards gay men more effectively than stereotypes and beliefs (Haddock et al., 1993). This pattern of results is likely because affect influences basic approach–avoidance reactions, whereas stereotypes and beliefs are more abstract and have less direct implications for behavioral reactions (Talaska et al., 2008).

In relation to the third research aim, the structural equation model revealed significant positive indirect effects of prejudice and support for removal of rights on discriminatory decision-making via employability concerns. Specifically, concerns about the employability of candidates living with HIV partially and fully mediated, respectively, the relationship between the emotional (i.e., prejudice) and behavioral (i.e., support for discriminatory policies) components of attitude and discriminatory decision-making in job promotion. The observed mediation aligns with previous research indicating that concerns regarding the lack of employability skills, arising from insufficient knowledge and understanding of HIV, represent significant barriers to hiring and retaining individuals living with HIV (Liu et al., 2012; Rao et al., 2008).

### 4.1 Strengths, Limitations, and Future Directions

It is crucial to acknowledge the strengths of this research. Firstly, the study was conducted with a representative sample of Dutch workers, enhancing the external validity and generalizability of the results obtained. Secondly, measures were developed in collaboration with community members to accurately capture job promotion discrimination against people living with HIV in the Netherlands, as existing validated measures were not sufficiently tailored to the HIV context. Thirdly, considering participants’ high social desirability tendencies concerning HIV-related topics, the inclusion of these tendencies as covariates in the analysis helped reducing potential bias in the parameter estimates. Moreover, the persistence of statistically significant effects even when adjusting for social desirability underscores the robustness of the findings, which could be even stronger in reality. Lastly, to the best of the authors’ knowledge, this dataset is the first to focus on job promotion discrimination against professionals living with HIV in the Netherlands, providing crucial insights into the attitudes of the current Dutch workforce.

Despite these strengths, there are some limitations to consider. The self-reported nature of the data may not accurately reflect individuals’ actual behavior regarding job promotion discrimination against candidates living with HIV. Additionally, while employability scales are available in the literature, their insufficient focus on HIV-specific issues (e.g., perceived absenteeism, perceived international health risks) required the creation of a novel measure based on experiences shared by the community. Lastly, the potential for measurement error in the chosen outcome (i.e., job promotion) cannot be overlooked, as it may have inadvertently led participants to express discriminatory decisions towards certain chronic conditions, even if unintended. This measurement error is likely the reason why the variables in the model accounted for only 25% of the variance in the outcome.

Given these limitations, future research will benefit from employing alternative outcome measures (e.g., archival data about job promotions) and research designs (e.g., longitudinal designs, audit studies) to further investigate job promotion discrimination against PLHIV and to assess the replicability of the findings of this study. Additionally, future studies should explore the role of intersectional stigma in shaping the career trajectories of professionals living with HIV. For instance, given the substantial body of literature on the relationship between leadership and gender stereotypes (Galsanjigmed & Sekiguchi, 2023), additional research could dive into how gender discrimination interacts with HIV stigma in decisions regarding promotion to leadership positions. Finally, more research is needed to explore the underlying dimensions of people’s perceptions of chronic health conditions and the differences among them. This could involve investigating factors such as perceived contagiousness, perceived responsibility, and the perceived severity of symptoms (Stutterheim et al., 2012). The multidimensional unfolding approach used in this study indeed provided an initial exploration of these dimensions, but more systematic analyses are warranted. It is important to note that the axes of the multidimensional unfolding plot do not have a predefined meaning; rather, their interpretation is at the discretion of the researcher and may vary depending on axis rotation. Therefore, multidimensional unfolding should be seen as a suggestive and exploratory tool for identifying clusters or patterns in the data, rather than as a definitive representation. By further exploring these dimensions, researchers can develop a more comprehensive understanding of the relationship between perceptions of illness and job promotion discrimination.

### 4.2 Implications

In line with Thornicroft and colleagues’ (2007) conceptualization of ignorance as a key driver of stigma, the current research highlight the need to address knowledge gaps within the Dutch population regarding HIV as a chronic condition, rather than a terminal one, and its non-disruptive impact on job performance. Prior studies have indeed shown that, although individuals living with HIV may have a lower employment rate (e.g., Legarth et al., 2014), employed professionals living with HIV exhibit negligible differences in health impairment and job productivity compared to the general population (Verbooy et al., 2018). Practical recommendations include fostering organizational practices that disseminate accurate, updated, and relevant information on HIV and comorbidities to employees, HR professionals, and managers (UNAIDS, 2020), with the overarching goal of changing potentially negative and unfounded attitudes towards colleagues living with HIV (Lau et al., 2005). Such stigma-reduction interventions could provide evidence that individuals living with HIV can lead long and healthy lives with antiretroviral medication (Stutterheim et al., 2008), while also highlighting the role of distress and discrimination experienced after the diagnosis, rather than the illness itself, in compromising the psychophysical health of individuals living with HIV (Thornicroft et al., 2007; Wagener et al., 2018). In this way, an HIV-stigma-free workplace that ensures everyone’s right to work (UNAIDS, 2020) and guarantees full and productive employment and decent work for all (UN General Assembly, 2015) can be achieved.

## 5. Conclusion

Overall, the findings of the current study expand the understanding of HIV workplace stigma by highlighting its impact on the promotion of people living with HIV to leadership positions. The results show that stigmatizing attitudes and inaccurate beliefs about the ability of professionals living with HIV to successfully carry out their job are prevalent in the Netherlands and significantly affect their career advancement opportunities. Continued investigation into how HIV discrimination shapes the career trajectories of people living with HIV, and how it intersects with other forms of discrimination, is necessary. Addressing these attitudes through education and organizational policy changes is crucial to creating inclusive workplaces that support the professional growth of all employees.

## Data Availability

Data used will be made available upon reasonable request to the corresponding author

