## Supplementary material for "“Let’s better not choose someone with HIV”: Dutch professionals’ attitudes and job promotion decisions regarding candidates living with HIV and the mediating role of employability concerns"

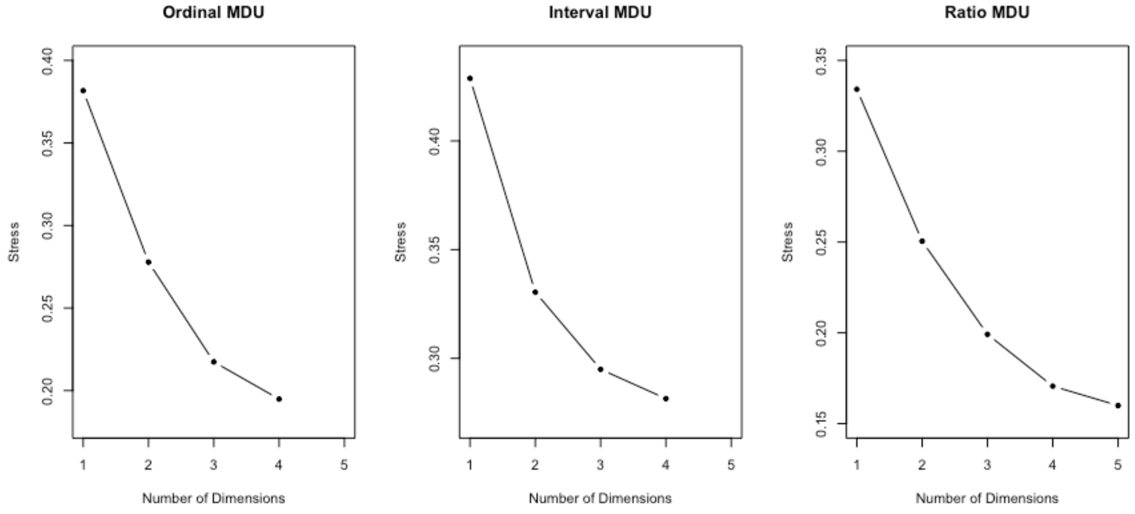


**Figure S1**. Scree plots for ordinal MDU, interval MDU, and ratio MDU solutions

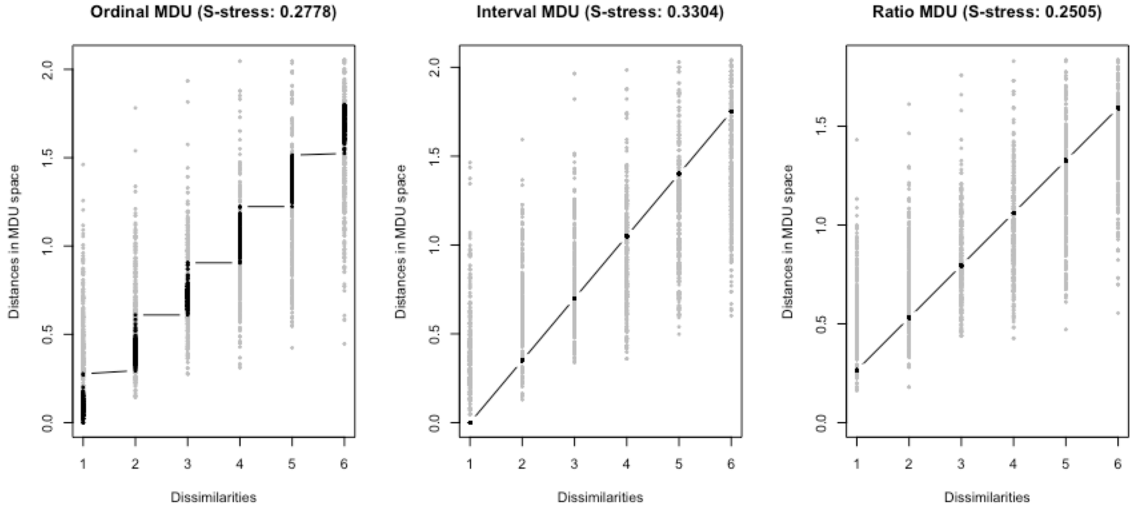


**Figure S2.** Shepard diagrams for ordinal MDU, interval MDU, and ratio MDU solutions

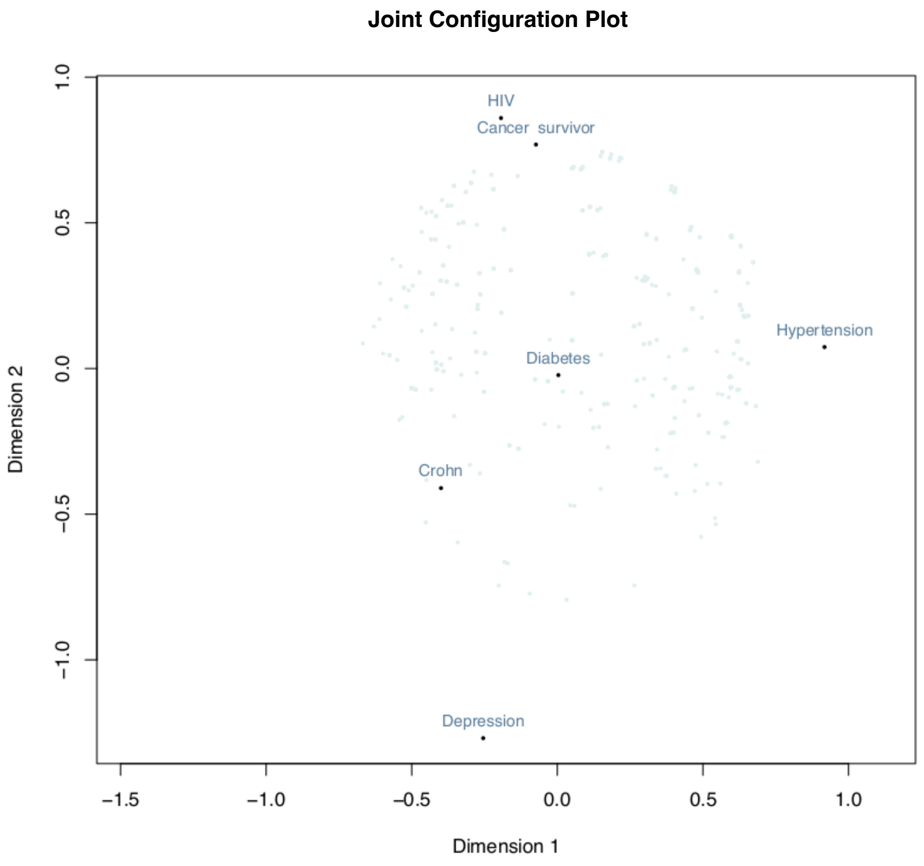


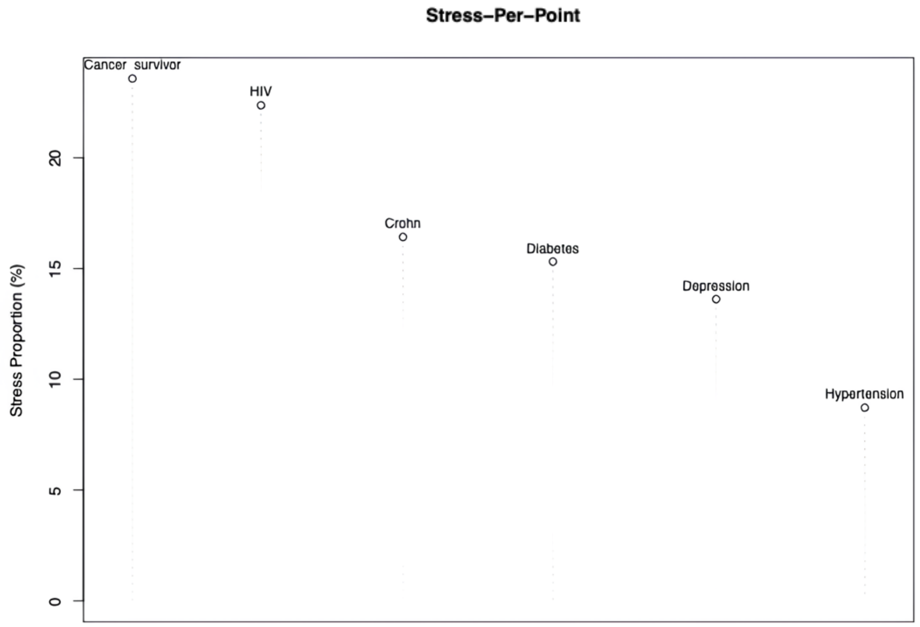


**Figure S3.** Top: Two-dimensional unfolding ordinal solution plot of individuals (light dots) and objects (dark dots). Bottom: Stress Per Point diagram illustrating the contribution of each object to the overall stress of the solution.
